# Risk factors for workplace bullying, severe psychological distress, and suicidal ideation during the COVID-19 pandemic: a nationwide internet survey for the general working population in Japan

**DOI:** 10.1101/2021.11.18.21266501

**Authors:** Kanami Tsuno, Takahiro Tabuchi

## Abstract

**Objectives:** The pandemic of the new coronavirus disease (COVID-19) has created a challenging environment for workers. This study aimed to investigate the risk factors for workplace bullying and mental health outcomes during the pandemic among workers.

**Methods:** We conducted a nationwide online cross-sectional survey from August to September 2020 in Japan (N = 16,384). Workplace bullying was measured by one item from the Brief Job Stress Questionnaire; severe psychological distress (SPD) by K6 (≥13); and suicidal ideation by one item. Prevalence ratios were calculated by Poisson regression analyses adjusting for potential confounders such as gender, age, occupational characteristics, and a prior history of depression.

**Results:** Overall, 15% of workers experienced workplace bullying, 9% had SPD, and 12% had suicidal ideation during the second and third wave of the COVID-19 pandemic in Japan. The results of this study showed men, executives, managers, and permanent employees had a higher risk of bullying compared to women or part-time workers.

Increased physical and psychological demands were common risk factors for bullying, SPD, and suicidal ideation. Newly starting working from home was a significant predictor for adverse mental health outcomes, however, it was found to be a preventive factor against workplace bullying.

**Conclusions:** The results of this study found different high-risk groups for bullying or mental health during the pandemic. When intervening to decrease workplace bullying or mental health problems, we should focus on not only previously reported vulnerable workers but also workers who experienced a change of their working styles or job demands.

**Key messages:** *What is already known about this subject?:*

- Workplace bullying is one of the severe job stressors in the workplace that cause mental health problems.
- Health care workers, less-educated workers, and non-regular female workers have been reported to have greater psychological distress during the COVID-19 pandemic.

*What are the new findings?:*

- About 15% of workers experienced workplace bullying, 9% had SPD, and 12% had suicidal ideation during the pandemic in Japan.
- Men, executives, managers, and permanent workers had a higher risk of bullying compared to women or part-time workers.
- Increased physical or psychological demands were common risk factors for bullying, SPD, and suicidal ideation.
- While newly starting working from home was a preventive factor against workplace bullying, it was found to be a significant risk factor for adverse mental health outcomes.

*How might this impact on policy or clinical practice in the foreseeable future?:*

- The results of this study indicate a different pattern of high-risk groups for bullying or mental health during the pandemic.
- When intervening to decrease workplace bullying or mental health problems, we should focus on not only previously reported vulnerable workers but also workers who experienced a change of their working styles or job demands.

## INTRODUCTION

Workplace bullying is a form of repeated negative acts and one of the severe job stressors in the workplace. Although the prevalence of workplace bullying during the coronavirus disease 2019 (COVID-19) pandemic is unknown, a global prevalence before the pandemic was reported as 14.6% in the meta-analysis.^1^

Previous studies clearly show workplace bullying has a severe adverse effect on workers’ mental health. For example, longitudinal associations between workplace bullying and depression,^2^ post-traumatic stress disorder,^3^ and suicidal ideation^4^ have been reported in the systematic review or meta-analysis. Moreover, mental health problems are not only “outcomes” of workplace bullying, but also “antecedents” of workplace bullying. The meta-analysis studies on the association between workplace bullying and mental health have consistently reported that baseline mental health problems were associated with an increased risk of exposing workplace bullying.^2^ ^5^ Thus, when investigating the association between bullying and mental health, a reversed effect should be taken into consideration.

Women and younger workers were more likely to experience bullying,^6 7^ although the results on the association between age and bullying were found to be inconsistent in the recent systematic review.^6^ Low socioeconomic status (SES), measured by education, income, and occupation, was also reported as a risk factor for workplace bullying.^8^ This is probably because lower SES workers tend to be in a lower position in the organization than higher SES workers, as shown by the report that workers with a higher position such as managers were less likely but unskilled workers were more likely to expose to bullying.^9^ Nevertheless, only one previous study for workplace bullying has focused on income,^8^ a few studies on education,^8 10 11^ and most other studies only investigated occupation as risk factors of workplace bullying.^6^ In addition to that low SES workers tend to have unstable working conditions, an economical shrinking during the COVID-19 pandemic may worsen their surroundings and increase their vulnerability.^12^ More research on workplace bullying needs to focus on the most disadvantaged workers in societies.

Although high job demands are associated with exposure to workplace bullying,^6 13–15^ no study investigated an association between increased job demands or new working style—working from home—and exposure to workplace bullying. Under the COVID-19 pandemic, especially essential workers, including health care workers, have experienced excessive overload physically and psychologically and have developed psychological distress or burnout.^16 17^ In contrast, non-essential workers such as white-collar workers started to work from home and the number of people working from home increased during the pandemic.^18^ These changes in working styles or the workplaces also cause workplace bullying because any kind of change makes workers feel stressful and stressful working environments increase bullying behaviors.^7 19 20^

An association between working from home and adverse mental health is not fully investigated. During the pandemic, suicide rates increased from 2019 to 2020 in Japan, which was the first time in the decade.^21^ Determinants of the increase in suicide rates have not been fully investigated yet but a recent study reported an increase in social isolation was associated with suicidal ideation in the general population during the pandemic.^22^ In general, workers are less likely to be isolated compared to unemployed people because they tend to have daily opportunities to meet and talk with supervisors or co-workers in the workplace. However, workers who work from home may be in a different situation; they have less co-worker support than workers who work at the office, while working from home itself has a positive impact on workers’ work-life balance.^23^ A recent large-scale study in an information company with over 60,000 employees during the pandemic showed that working from home has negative effects such as decreasing synchronous communication between workers and decreasing bridges between disparate departments in the company.^24^

Various risk factors of mental health problems among workers have been reported during the COVID-19 pandemic. For example, health care workers,^17 25^ less-educated workers,^26^ and non-regular female workers^27^ were more likely to have greater psychological distress. However, studies focusing on the general working population from various industries are scarce, since a majority of studies have focused on only health care workers.^25^

The aim of this study was therefore to identify the potential risk factors of workplace bullying, severe psychological distress (SPD), and suicidal ideation during the pandemic, such as gender, age, SES, job demands, and working from home, by a nationwide internet survey for the general working population in Japan.

## METHODS

### Data

We used the baseline cross-sectional data of an ongoing web-based national-representative longitudinal study, the Japan “COVID-19 and Society” Internet Survey (JACSIS) study. The baseline survey was conducted in August and September 2020. The survey requests were sent by the research agency (Rakuten Insight, Inc., Tokyo, Japan) to the 224,389 panelists who were selected by each gender, age, and prefecture category using simple random sampling. Once the target number of participants (N = 28,000) answered the questionnaire, the recruitment process stopped, resulting in the participation rate for the survey being 12.5% (28,000 of 224,389). The details of the study protocol are described elsewhere.^28^

To validate data quality, we excluded respondents showing discrepancies and/or artificial/unnatural responses.^29^ Three question items of “Please choose the second from the bottom”, “choosing positive in all of a set of questions for using drugs” and “choosing positive in all of a set of questions for having chronic diseases” were used to detect any discrepancies. We excluded these respondents with discrepancies or artificial/unnatural responses (n=2518, remaining n=25,482).

## Measurements

### Risk factors

Our exposures variables of interest were respondents’ demographics variables including gender,^6^ age,^7^ living areas, and marital status (having a partner/spouse), SES,^8^ occupational characteristics, and current working situation. The SES variables included education (high school or below, junior college/vocational school, and university of above)^8^, annual household income during the previous year (JPY1.99 million, JPY2.00-3.99 million, JPY4.00-5.99 million, JPY6.00-7.99 million, JPY8.00-9.99 million, JPY10 million, and unknown), and occupation/employment status (executive, self-employee/individual business owner, family business assistance, manager, permanent worker, dispatched worker, contract worker, and part-time worker).

Occupational characteristics included industry, office size, and job type (desk work, jobs that work with people [sales staff, in-store salesperson, etc.], and jobs that require physical strength [delivery staff, care staff, etc.]). To assess their current working situations, we asked whether the respondents experienced working from home or experienced increased physical or psychological demands during the COVID-19 pandemic. Weekly working hours during the previous month were also assessed as categorical variables.

Finally, a prior history of depression and other mental illnesses were assessed since baseline mental health problems were associated with an increased risk of exposure to workplace bullying.^2^ ^5^

### Workplace bullying

Workplace bullying was assessed by a self-labeling method, using a sub-scale of the New Brief Job Stress Questionnaire.^8^ First, respondents were asked whether they experienced bullying during the six months, using a single-item “Have you been bullied in your workplace during the six months (since April 2020)?” The respondents who chose “yes” were defined as “victims”. In the survey, we did not present a definition of bullying to respondents due to limitations of space. In addition to the abovementioned question, respondents were also asked whether they had witnessed bullying in their workplace during the six months.

### Mental health outcomes

Severe psychological distress was measured by the K6.^30^ The K6 consists of six items and assesses how frequently respondents have experienced symptoms of psychological distress during the past 30 days (“0 = never”, “1 = rarely”, “2 = sometimes”, “3 = often” or “4 = always”). In this study, a cut-off score of 13 was used for defining severe psychological distress (SPD).^31^

Suicidal ideation was assessed by one question “Have you ever wished you were dead, since April 2020?” The response options were “1 = yes, for the first time”, “2 = yes, but I had it since before April 2020” or “3 = never experienced it.” Answering “yes” was defined as having suicidal ideation.

## Statistical analyses

We used a Poisson regression analysis to examine the relationship between risk factors and workplace bullying. Prevalence ratios (PRs) and 95% confidence intervals (CIs) were calculated adjusting for individual characteristics (gender, age, having a partner, and living area) and SES (education, household income, and employment status) (Model 1), occupational characteristics (industry, office size, and job type) (Model 2), and all variables including current working situation (started to work from home after the pandemic, increased physical demands, increased psychological demands, and weekly working hours during the previous month), and a prior history of depression (Model 3). To examine the relationship between workplace bullying and mental health outcomes, we also conducted another Poisson regression analysis. In these analyses, PRs and 95% CIs were calculated adjusting for individual characteristics, SES, occupational characteristics (Model 1), and a prior history of depression (Model 2). Finally, we conducted a Poisson regression analysis stratified by gender. In this analysis, the prevalence ratios of two mental health outcomes were calculated by adjusting individual characteristics, SES, occupational characteristics, workplace bullying, and a prior history of depression. The 2-tailed p-value for statistical significance to see the differences among each social indicator was set at 0.05. All analyses were conducted using SPSS 27.0 for Windows. No missing values exist in this data because all questions were required to answer.

## RESULTS

### Characteristics of participants

Table 1 shows the characteristics of the participants. Among 25,482 respondents, we analyzed for 16,384 workers in this study after excluding students, retired persons, full-time housewives/househusbands, and those who did not work at the survey. The average age of the participants was 45.7 (standard deviation: 13.8) years old. The majority of the participants were men, were 45-54 years old, had a partner/spouse, and lived in prefectures under special precautions during the first COVID-19 state of emergency in Japan (April-May 2020). Regarding SES variables, the majority of the participants have graduated university or graduate school, earned 4.00-5.99 million Japanese yen during the previous year as a household income, and worked as permanent workers. In terms of occupational characteristics, the majority of the participants were working in the manufacturing industry, working at a small office with 5-29 employees or a large office with more than 1,000 employees. Their job type was mainly desk work. In terms of current working situations, only 8% of the participants started to work from home after the pandemic but approximately 20% were working from home since before the pandemic, which means in total about 30% of participants experienced working from home during the COVID-19 pandemic. Although most of the participants worked 40-44 hours/week during the past month, 6% worked over 60 hours/week. Overall, 21% experienced increased physical demands and 33% experienced increased psychological demands during the pandemic. About 4% of the participants had depression or other mental illness at the time of the survey and 6% or 4% had a prior history of depression or other mental illness, respectively.

**Table 1.** Characteristics of the participants (N=16,384)

| **Individual characteristics** | **n** | **%** |
| --- | --- | --- |
| Gender |  |  |
| Men | 9595 | 58.6 |
| Women | 6789 | 41.4 |
| Age |  |  |
| Under 24 | 1024 | 6.3 |
| 25-34 | 2964 | 18.1 |
| 35-44 | 3673 | 22.4 |
| 45-54 | 4146 | 25.3 |
| 55-64 | 2914 | 17.8 |
| Over 65 | 1663 | 10.2 |
| Having a partner/spouse |  |  |
| Yes | 9633 | 58.8 |
| No | 6751 | 41.2 |
| Living area |  |  |
| Prefectures under special precautions | 10246 | 62.5 |
| Others | 6138 | 37.5 |
| **Socioeconomic status (SES)** |  |  |
| Education |  |  |
| High school or below | 4167 | 25.4 |
| Junior college/vocational school | 3658 | 22.3 |
| University or above | 8559 | 52.2 |
| Annual household income during the previous year (million JPY) |  |  |
| 1.99 or less | 987 | 6.0 |
| 2.00-3.99 | 3053 | 18.6 |
| 4.00-5.99 | 3469 | 21.2 |
| 6.00-7.99 | 2481 | 15.1 |
| 8.00-9.99 | 1744 | 10.6 |
| 10.00 or more | 1998 | 12.2 |
| Unknown | 2652 | 16.2 |
| Occupation/employment status |  |  |
| Executive | 927 | 5.7 |
| Self-employee/individual business owner | 1548 | 9.4 |
| Family business assistance | 210 | 1.3 |
| Manager | 2014 | 12.3 |
| Permanent worker (non-manager) | 7201 | 44.0 |
| Dispatched worker | 366 | 2.2 |
| Contract worker | 1062 | 6.5 |
| Part-time worker | 3056 | 18.7 |
| **Occupational characteristics** |  |  |
| Industry |  |  |
| Public administration | 1065 | 6.5 |
| Agriculture, forestry, and fishing | 181 | 1.1 |
| Construction | 908 | 5.5 |
| Manufacturing | 2748 | 16.8 |
| Electricity, gas, and water supply | 235 | 1.4 |
| Telecommunication | 844 | 5.2 |
| Transport | 684 | 4.2 |
| Wholesale | 571 | 3.5 |
| Retail trade | 1269 | 7.7 |
| Finance | 423 | 2.6 |
| Insurance | 288 | 1.8 |
| Real estate | 396 | 2.4 |
| Restaurants | 508 | 3.1 |
| Hotels | 151 | 0.9 |
| Healthcare | 1201 | 7.3 |
| Welfare | 704 | 4.3 |
| Education and learning assistance | 853 | 5.2 |
| Others | 3355 | 20.5 |
| Office size |  |  |
| 1-4 | 2379 | 14.5 |
| 5-29 | 3241 | 19.8 |
| 30-49 | 1161 | 7.1 |
| 50-99 | 1625 | 9.9 |
| 100-299 | 2145 | 13.1 |
| 300-499 | 999 | 6.1 |
| 500-999 | 1065 | 6.5 |
| Over 1,000 | 3158 | 19.3 |
| Civil service | 611 | 3.7 |
| Job types |  |  |
| Desk work | 7944 | 48.5 |
| Jobs that work with people | 4024 | 24.6 |
| Jobs that require physical strength | 4416 | 27.0 |

**Table 1 (cont.).** Characteristics of the participants (N=16,384)
|  | n | % |
| --- | --- | --- |
| **Current working situation** |  |  |
| Started to work from home after the pandemic |  |  |
| Yes | 1382 | 8.4 |
| No | 15002 | 91.6 |
| Working from home since before the pandemic |  |  |
| Yes | 2964 | 18.1 |
| No | 13420 | 81.9 |
| Increase in physical demands |  |  |
| Yes | 3389 | 20.7 |
| No | 12995 | 79.3 |
| Increase in psychological demands |  |  |
| Yes | 5421 | 33.1 |
| No | 10963 | 66.9 |
| Weekly working hours during the previous month |  |  |
| Less than 20 hours/week | 2688 | 16.4 |
| 20-29 hours/week | 1821 | 11.1 |
| 30-39 hours/week | 3103 | 18.9 |
| 40-44 hours/week | 4676 | 28.5 |
| 45-49 hours/week | 1900 | 11.6 |
| 50-59 hours/week | 1265 | 7.7 |
| Over 60 hours/week | 931 | 5.7 |
| **History of psychiatric disorders** |  |  |
| Having mental illness |  |  |
| Depression |  |  |
| Never | 14782 | 90.2 |
| Past | 989 | 6.0 |
| Current | 613 | 3.7 |
| Other mental illness |  |  |
| Never | 15203 | 92.8 |
| Past | 611 | 3.7 |
| Current | 570 | 3.5 |
| Exposure to workplace bullying |  |  |
| Yes | 2441 | 14.9 |
| No | 13943 | 85.1 |
| Witnessed workplace bullying |  |  |
| Yes | 2940 | 17.9 |
| No | 13444 | 82.1 |
| SPD (K6 ≥ 13) | | |
| Yes | 1442 | 8.8 |
| No | 14942 | 91.2 |
| Having suicidal ideation |  |  |
| Yes | 1890 | 11.5 |
| No | 14494 | 88.5 |

SPD: severe psychological distress.

Overall, 15% of the participants experienced workplace bullying during the past 6 months and 18% witnessed bullying at their workplaces during the past 6 months. About 9% had SPD during the past 30 days and 12% had suicidal ideation during the past 6 months.

### Risk factors for exposure to workplace bullying

Table 2 shows the results of the Poisson regression analysis, which calculated the PRs for workplace bullying. The significant risk factors of workplace bullying were gender (men), younger age, lower household income (1.99-3.99 million Japanese yen), occupation (executive, manager, permanent employee, and contract employee), larger office size, increased physical or psychological demands, and current or a prior history of depression or other mental illness. On the other hand, those who started to work from home after the pandemic or worked 30-49 hours/week had a lower risk of exposure to workplace bullying..

**Table 2.**
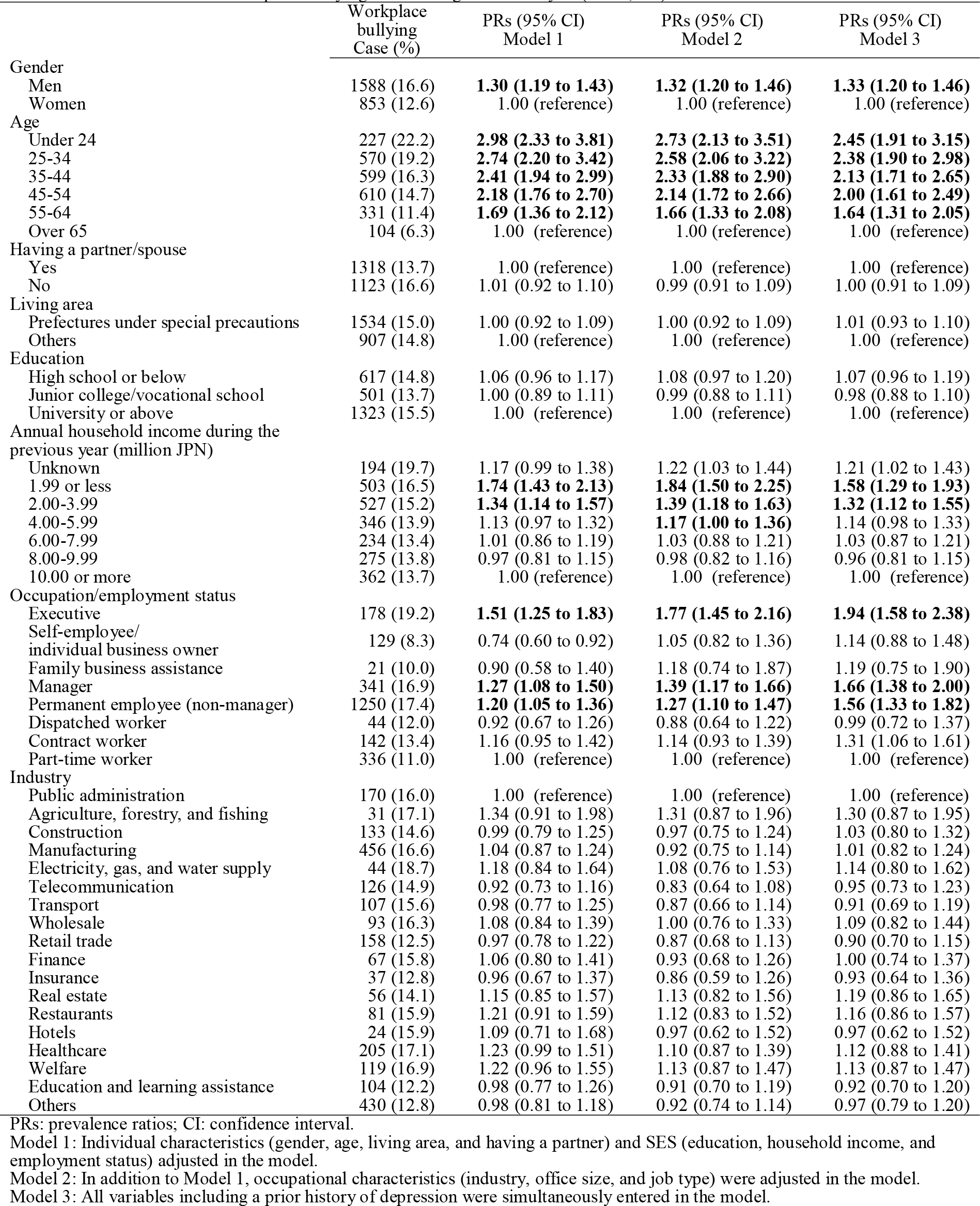
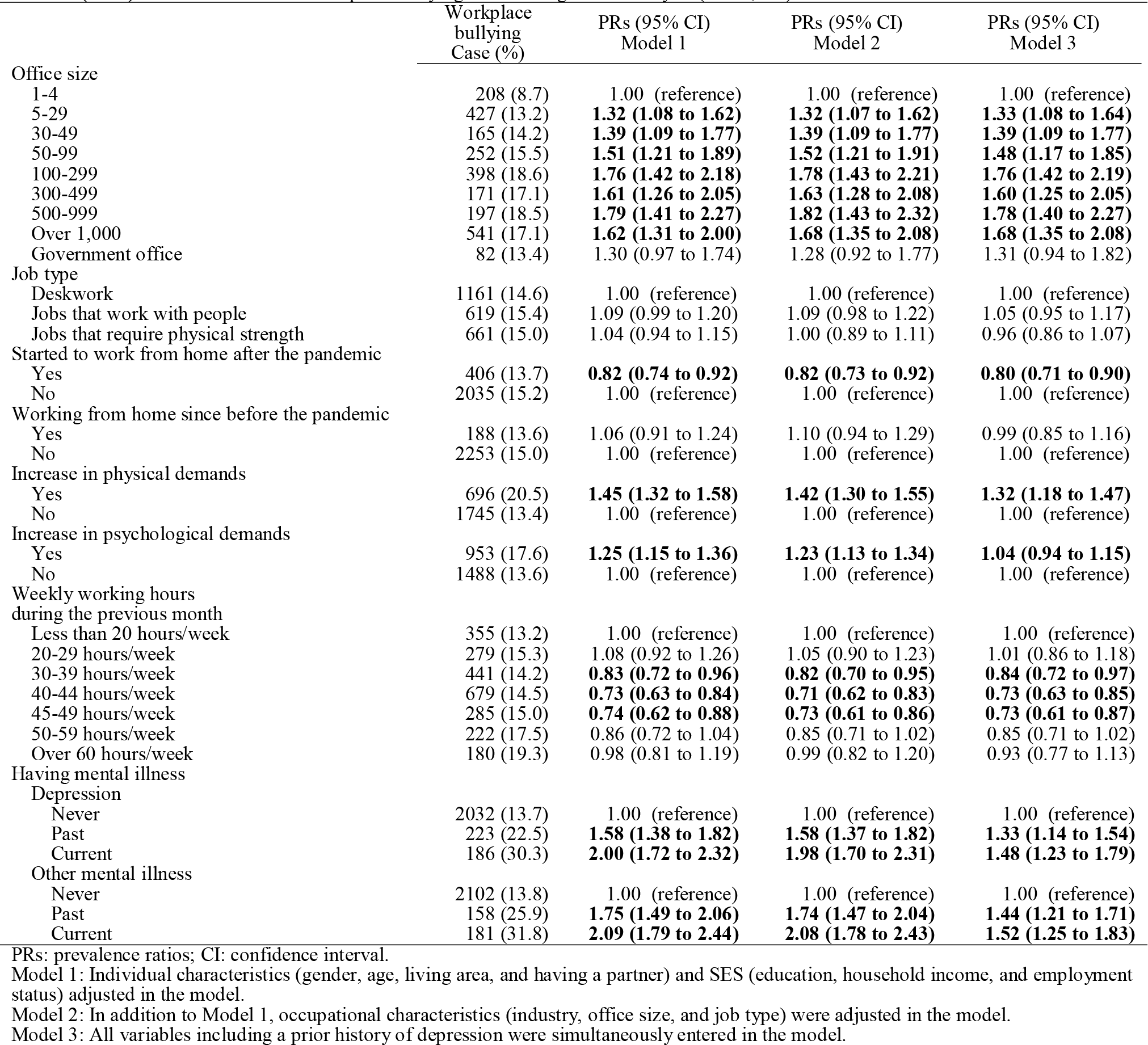
Prevalence ratios of workplace bullying: Poisson regression analysis (N=16,384)

### Association between workplace bullying and mental health outcomes

Exposure to workplace bullying was significantly associated with SPD and suicidal ideation (PR for SPD: 2.28 [95%CI: 2.53-3.18]; PR for suicidal ideation: 1.92 [1.73-2.13]), after adjusting for individual characteristics, SES, occupational characteristics, and a prior history of depression (Model 2 in Table 3). Although larger PRs were observed for exposure to workplace bullying, witness to bullying was also significantly associated with SPD and suicidal ideation in Model 2 (PR for SPD: 1.90 95%CI: 1.58-2.28]; PR for suicidal ideation: 1.39 [1.17-1.65]). When stratified by gender, men who experienced workplace bullying had higher PRs for both SPD and suicidal ideation than women (PR for SPD in men: 3.20 [95%CI: 2.74-3.73] vs in women 2.39 [2.00-2.86]; PR for suicidal ideation in men: 1.95 [1.70-2.25] vs in women 1.87 [1.59-2.19]).

**Table 3.** Workplace bullying and mental health outcomes: Poisson regression analysis

|  | SPD |  |  | Suicidal ideation |  |  |
| --- | --- | --- | --- | --- | --- | --- |
|  | Case/<br>All<br>(%) | PRs (95% CI)<br>Model 1 | PRs (95% CI)<br>Model 2 | Case/<br>All<br>(%) | PRs (95% CI)<br>Model 1 | PRs (95% CI)<br>Model 2 |
| **All (N = 16,384)** |  |  |  |  |  |  |
| Not exposed<br>nor witnessed | 761/<br>12,869<br>(5.9) | 1.00 (reference) | 1.00 (reference) | 1,182/<br>12,869<br>(9.2) | 1.00 (reference) | 1.00 (reference) |
| Not exposed<br>but witnessed | 135/<br>1,074<br>(12.6) | **2.01 (1.67 to 2.41)** | **1.90 (1.58 to 2.28)** | 150/<br>1,074<br>(14.0) | **1.48 (1.24 to 1.75)** | **1.39 (1.17 to 1.65)** |
| Exposed | 546/<br>2,441<br>(22.4) | **3.30 (2.95 to 3.69)** | **2.28 (2.53 to 3.18)** | 558/<br>2,441<br>(22.9) | **2.24 (2.02 to 2.48)** | **1.92 (1.73 to 2.13)** |
| **Men (N = 9,565)** |  |  |  |  |  |  |
| Not exposed<br>nor witnessed | 365/<br>7,361<br>(5.0) | 1.00 (reference) | 1.00 (reference) | 593/<br>7,361<br>(8.1) | 1.00 (reference) | 1.00 (reference) |
| Not exposed<br>but witnessed | 81/<br>646<br>(12.5) | **2.30 (1.80 to 2.94)** | **2.16 (1.69 to 2.76)** | 92/<br>646<br>(14.2) | **1.67 (1.34 to 2.09)** | **1.55 (1.24 to 1.93)** |
| Exposed | 357/<br>1,588<br>(22.5) | **3.69 (3.18 to 4.30)** | **3.20 (2.74 to 3.73)** | 340/<br>1,588<br>(21.4) | **2.27 (1.98 to 2.61)** | **1.95 (1.70 to 2.25)** |
| **Women (N = 6,789)** |  |  |  |  |  |  |
| Not exposed<br>nor witnessed | 396/<br>5,508<br>(7.2) | 1.00 (reference) | 1.00 (reference) | 589/<br>5,508<br>(10.7) | 1.00 (reference) | 1.00 (reference) |
| Not exposed<br>but witnessed | 54/<br>428<br>(12.6) | **1.72 (1.29 to 2.29)** | **1.66 (1.24 to 2.21)** | 58/<br>428<br>(13.6) | 1.26 (0.96 to 1.65) | 1.21 (0.92 to 1.59) |
| Exposed | 189/<br>853<br>(22.2) | **2.81 (2.36 to 3.36)** | **2.39 (2.00 to 2.86)** | 218/<br>6,789<br>(12.7) | **2.19 (1.87 to 2.57)** | **1.87 (1.59 to 2.19)** |

SPD: severe psychological distress; PRs: prevalence ratios; CI: confidence interval.
Model 1: Adjusted for individual characteristics (gender, age, living area, and having a partner), SES (education, household income, and employment status), and occupational characteristics (industry, office size, and job type).
Model 2: In addition to Model 1, a prior history of depression was adjusted.

### Other risk factors for mental health outcomes

In men, younger age, not having a partner, low household income (1.99 million Japanese yen), working from home since before the pandemic, increase in physical or psychological demands during the pandemic, and current or a prior history of depression were significantly and independently associated with both SPD and suicidal ideation in the workplace bullying adjusted model (Table 4). In women, similar trends were observed but two different results were found: manufacturing industry was significantly and negatively associated with SPD; permanent employment was also significantly and negatively associated with suicidal ideation, after adjusting for workplace bullying (Table 5).

**Table 4.** Risk factors for mental health outcomes among men (N = 9,565): Poisson regression analysis

|  | SPD |  |  |  | Suicidal ideation |  |  |
| --- | --- | --- | --- | --- | --- | --- | --- |
|  | All | Case | % | PRs (95% CI)† | Case | % | PRs (95% CI)† |
| Age |  |  |  |  |  |  |  |
| Under 24 | 512 | 94 | 18.4 | **1.78 (1.03 to 3.08)** | 119 | 23.2 | **3.73 (2.48 to 5.59)** |
| 25-34 | 1659 | 221 | 13.3 | **3.39 (2.02 to 5.68)** | 240 | 14.5 | **3.12 (2.13 to 4.56)** |
| 35-44 | 2112 | 212 | 10.0 | **4.12 (2.46 to 6.90)** | 269 | 12.7 | **2.97 (2.05 to 4.29)** |
| 45-54 | 2431 | 196 | 8.1 | **4.83 (2.87 to 8.14)** | 258 | 10.6 | **2.48 (1.72 to 3.59)** |
| 55-64 | 1816 | 63 | 3.5 | **4.76 (2.75 to 8.25)** | 103 | 5.7 | **1.54 (1.05 to 2.28)** |
| Over 65 | 1065 | 17 | 1.6 | 1.00 (reference) | 36 | 3.4 | 1.00 (reference) |
| Having a partner/spouse |  |  |  |  |  |  |  |
| Yes | 6093 | 369 | 6.1 | 1.00 (reference) | 464 | 7.6 | 1.00 (reference) |
| No | 3502 | 434 | 12.4 | **1.31 (1.12 to 1.54)** | 561 | 16.0 | **1.34 (1.16 to 1.54)** |
| Living area |  |  |  |  |  |  |  |
| Prefectures under special precautions | 6008 | 526 | 8.8 | 1.05 (0.90 to 1.22) | 662 | 11.0 | 1.03 (0.90 to 1.17) |
| Others | 3587 | 277 | 7.7 | 1.00 (reference) | 363 | 10.1 | 1.00 (reference) |
| Education |  |  |  |  |  |  |  |
| High school or below | 2246 | 179 | 8.0 | 0.94 (0.78 to 1.13) | 249 | 11.1 | 0.97 (0.83 to 1.14) |
| Junior college/vocational school | 1360 | 112 | 8.2 | 0.92 (0.74 to 1.14) | 158 | 11.6 | 1.02 (0.85 to 1.22) |
| University or above | 5989 | 512 | 8.5 | 1.00 (reference) | 618 | 10.3 | 1.00 (reference) |
| Annual household income during the previous year (million yen) |  |  |  |  |  |  |  |
| Unknown | 1241 | 66 | 5.3 | 0.76 (0.54 to 1.05) | 100 | 8.1 | 1.04 (0.77 to 1.40) |
| 1.99 or less | 450 | 86 | 19.1 | **1.57 (1.13 to 2.17)** | 117 | 26.0 | **2.08 (1.54 to 2.82)** |
| 2.00-3.99 | 1592 | 185 | 11.6 | 1.30 (0.99 to 1.72) | 234 | 14.7 | **1.57 (1.21 to 2.04)** |
| 4.00-5.99 | 2177 | 175 | 8.0 | 1.01 (0.77 to 1.32) | 241 | 11.1 | **1.38 (1.07 to 1.78)** |
| 6.00-7.99 | 1602 | 120 | 7.5 | 1.08 (0.81 to 1.42) | 140 | 8.7 | 1.23 (0.94 to 1.61) |
| 8.00-9.99 | 1164 | 80 | 6.9 | 0.98 (0.72 to 1.33) | 102 | 8.8 | 1.25 (0.94 to 1.66) |
| 10.00 or more | 1369 | 91 | 6.6 | 1.00 (reference) | 91 | 6.6 | 1.00 (reference) |
| Occupation/Employment status |  |  |  |  |  |  |  |
| Executive | 733 | 68 | 9.3 | 1.06 (0.73 to 1.55) | 77 | 10.5 | 0.92 (0.66 to 1.28) |
| Self-employee/individual business owner | 1144 | 79 | 6.9 | 1.37 (0.90 to 2.08) | 120 | 10.5 | 1.04 (0.73 to 1.47) |
| Family business assistance | 69 | 15 | 21.7 | **2.57 (1.40 to 4.74)** | 12 | 17.4 | 1.10 (0.58 to 2.08) |
| Manager | 1699 | 111 | 6.5 | 0.84 (0.59 to 1.20) | 133 | 7.8 | 0.80 (0.59 to 1.09) |
| Permanent worker (non-manager) | 4568 | 423 | 9.3 | 0.90 (0.66 to 1.22) | 516 | 11.3 | 0.84 (0.65 to 1.09) |
| Dispatched worker | 142 | 17 | 12.0 | 1.07 (0.61 to 1.88) | 27 | 19.0 | 1.19 (0.76 to 1.86) |
| Contract worker | 598 | 31 | 5.2 | 0.76 (0.48 to 1.19) | 51 | 8.5 | 0.81 (0.57 to 1.16) |
| Part-time worker | 642 | 59 | 9.2 | 1.00 (reference) | 89 | 13.9 | 1.00 (reference) |
| Industry |  |  |  |  |  |  |  |
| Public administration | 771 | 73 | 9.5 | 1.00 (reference) | 69 | 8.9 | 1.00 (reference) |
| Agriculture, forestry, and fishing | 134 | 19 | 14.2 | 1.32 (0.76 to 2.28) | 19 | 14.2 | 1.24 (0.72 to 2.14) |
| Construction | 623 | 37 | 5.9 | 0.75 (0.49 to 1.16) | 55 | 8.8 | 1.00 (0.68 to 1.48) |
| Manufacturing | 2069 | 182 | 8.8 | 1.03 (0.74 to 1.42) | 206 | 10.0 | 1.03 (0.75 to 1.41) |
| Electricity, gas, and water supply | 173 | 16 | 9.2 | 0.93 (0.53 to 1.65) | 24 | 13.9 | 1.31 (0.80 to 2.15) |
| Telecommunication | 649 | 50 | 7.7 | 0.88 (0.59 to 1.32) | 78 | 12.0 | 1.22 (0.85 to 1.76) |
| Transport | 523 | 38 | 7.3 | 0.81 (0.52 to 1.26) | 66 | 12.6 | 1.22 (0.83 to 1.79) |
| Wholesale | 376 | 33 | 8.8 | 1.09 (0.70 to 1.71) | 43 | 11.4 | 1.30 (0.86 to 1.96) |
| Retail trade | 538 | 39 | 7.2 | 0.93 (0.60 to 1.44) | 60 | 11.2 | 1.21 (0.82 to 1.80) |
| Finance | 239 | 12 | 5.0 | 0.66 (0.35 to 1.25) | 16 | 6.7 | 0.83 (0.47 to 1.47) |
| Insurance | 139 | 11 | 7.9 | 1.09 (0.56 to 2.11) | 10 | 7.2 | 0.96 (0.48 to 1.90) |
| Real estate | 261 | 16 | 6.1 | 0.97 (0.54 to 1.72) | 24 | 9.2 | 1.30 (0.79 to 2.13) |
| Restaurants | 170 | 23 | 13.5 | 1.29 (0.76 to 2.16) | 26 | 15.3 | 1.25 (0.76 to 2.05) |
| Hotels | 68 | 7 | 10.3 | 0.95 (0.42 to 2.13) | 9 | 13.2 | 1.07 (0.52 to 2.21) |
| Healthcare | 414 | 39 | 9.4 | 1.08 (0.70 to 1.65) | 43 | 10.4 | 1.10 (0.72 to 1.66) |
| Welfare | 270 | 24 | 8.9 | 0.97 (0.59 to 1.60) | 32 | 11.9 | 1.09 (0.69 to 1.71) |
| Education and learning assistance | 402 | 35 | 8.7 | 0.97 (0.63 to 1.48) | 44 | 10.9 | 1.19 (0.80 to 1.78) |
| Others | 1776 | 149 | 8.4 | 1.02 (0.73 to 1.42) | 201 | 11.3 | 1.15 (0.83 to 1.58) |
| Office size |  |  |  |  |  |  |  |
| 1-4 | 1462 | 94 | 6.4 | 1.00 (reference) | 154 | 10.5 | 1.00 (reference) |
| 5-29 | 1569 | 121 | 7.7 | 1.26 (0.89 to 1.80) | 170 | 10.8 | 0.99 (0.74 to 1.33) |
| 30-49 | 615 | 46 | 7.5 | 1.22 (0.79 to 1.88) | 55 | 8.9 | 0.82 (0.57 to 1.20) |
| 50-99 | 892 | 89 | 10.0 | 1.31 (0.89 to 1.93) | 104 | 11.7 | 0.94 (0.68 to 1.31) |
| 100-299 | 1280 | 122 | 9.5 | 1.28 (0.88 to 1.86) | 151 | 11.8 | 0.96 (0.70 to 1.32) |
| 300-499 | 602 | 60 | 10.0 | 1.31 (0.86 to 2.00) | 74 | 12.3 | 1.01 (0.71 to 1.44) |
| 500-999 | 640 | 62 | 9.7 | 1.38 (0.91 to 2.11) | 73 | 11.4 | 1.04 (0.73 to 1.49) |
| Over 1,000 | 2094 | 169 | 8.1 | 1.23 (0.85 to 1.78) | 210 | 10.0 | 0.95 (0.70 to 1.30) |
| Government office | 441 | 40 | 9.1 | 1.41 (0.86 to 2.29) | 34 | 7.7 | 0.81 (0.51 to 1.31) |

SPD: severe psychological distress; PRs: prevalence ratios; CI: confidence interval.
† Individual characteristics (gender, age, living area, and having a partner), SES (education, household income, and employment status), occupational characteristics (industry, office size, and job type), workplace bullying, and a prior history of depression adjusted.

**Table 4 (cont.). Risk factors for mental health outcomes among men (N = 9,565): Poisson regression analysis**
|  | SPD |  |  |  | Suicidal ideation |
| --- | --- | --- | --- | --- | --- |

|  | All | Case | % | PRs (95% CI)† | Case | % | PRs (95% CI)† |
| --- | --- | --- | --- | --- | --- | --- | --- |
| Job type |  |  |  |  |  |  |  |
| Deskwork | 4795 | 395 | 8.2 | 1.00 (reference) | 492 | 10.3 | 1.00 (reference) |
| Jobs that work with people | 2145 | 200 | 9.3 | 1.02 (0.85 to 1.22) | 233 | 10.9 | 0.94 (0.79 to 1.11) |
| Jobs that require physical strength | 2655 | 208 | 7.8 | 0.91 (0.75 to 1.10) | 300 | 11.3 | 1.00 (0.85 to 1.18) |
| Started to work from home after the pandemic |  |  |  |  |  |  |  |
| Yes | 1973 | 178 | 9.0 | 1.15 (0.96 to 1.38) | 223 | 11.3 | **1.19 (1.01 to 1.40)** |
| No | 7622 | 625 | 8.2 | 1.00 (reference) | 802 | 10.5 | 1.00 (reference) |
| Working from home since before the pandemic |  |  |  |  |  |  |  |
| Yes | 888 | 112 | 12.6 | **1.49 (1.20 to 1.85)** | 140 | 15.8 | **1.45 (1.20 to 1.76)** |
| No | 8707 | 691 | 7.9 | 1.00 (reference) | 885 | 10.2 | 1.00 (reference) |
| Increase in physical demands |  |  |  |  |  |  |  |
| Yes | 1813 | 339 | 18.7 | **2.17 (1.87 to 2.51)** | 378 | 20.8 | **1.87 (1.64 to 2.14)** |
| No | 7782 | 464 | 6.0 | 1.00 (reference) | 647 | 8.3 | 1.00 (reference) |
| Increase in psychological demands |  |  |  |  |  |  |  |
| Yes | 2930 | 429 | 14.6 | **2.07 (1.80 to 2.39)** | 549 | 18.7 | **2.24 (1.97 to 2.54)** |
| No | 6665 | 374 | 5.6 | 1.00 (reference) | 476 | 7.1 | 1.00 (reference) |
| Weekly working hours during the previous month |  |  |  |  |  |  |  |
| Less than 20 hours/week | 943 | 90 | 9.5 | 1.00 (reference) | 120 | 12.7 | 1.00 (reference) |
| 20-29 hours/week | 747 | 101 | 13.5 | 1.14 (0.85 to 1.53) | 134 | 17.9 | 1.25 (0.97 to 1.60) |
| 30-39 hours/week | 1686 | 124 | 7.4 | 0.89 (0.67 to 1.18) | 175 | 10.4 | 0.94 (0.74 to 1.21) |
| 40-44 hours/week | 3040 | 216 | 7.1 | 0.87 (0.66 to 1.14) | 283 | 9.3 | 0.84 (0.66 to 1.06) |
| 45-49 hours/week | 1416 | 82 | 5.8 | **0.71 (0.51 to 0.97)** | 117 | 8.3 | 0.76 (0.58 to 1.01) |
| 50-59 hours/week | 1009 | 97 | 9.6 | 1.09 (0.80 to 1.49) | 103 | 10.2 | 0.92 (0.69 to 1.21) |
| Over 60 hours/week | 754 | 93 | 12.3 | 1.24 (0.91 to 1.68) | 93 | 12.3 | 0.99 (0.74 to 1.31) |
| History of depression |  |  |  |  |  |  |  |
| Never | 8644 | 544 | 6.3 | 1.00 (reference) | 706 | 8.2 | 1.00 (reference) |
| Past | 559 | 100 | 17.9 | **2.14 (1.72 to 2.65)** | 148 | 26.5 | **2.70 (2.26 to 3.24)** |
| Current | 392 | 159 | 40.6 | **4.00 (3.32 to 4.83)** | 171 | 43.6 | **3.69 (3.10 to 4.41)** |

SPD: severe psychological distress; PRs: prevalence ratios; CI: confidence interval.
† Individual characteristics (gender, age, living area, and having a partner), SES (education, household income, and employment status), occupational characteristics (industry, office size, and job type), workplace bullying, and a prior history of depression adjusted.

**Table 5.**
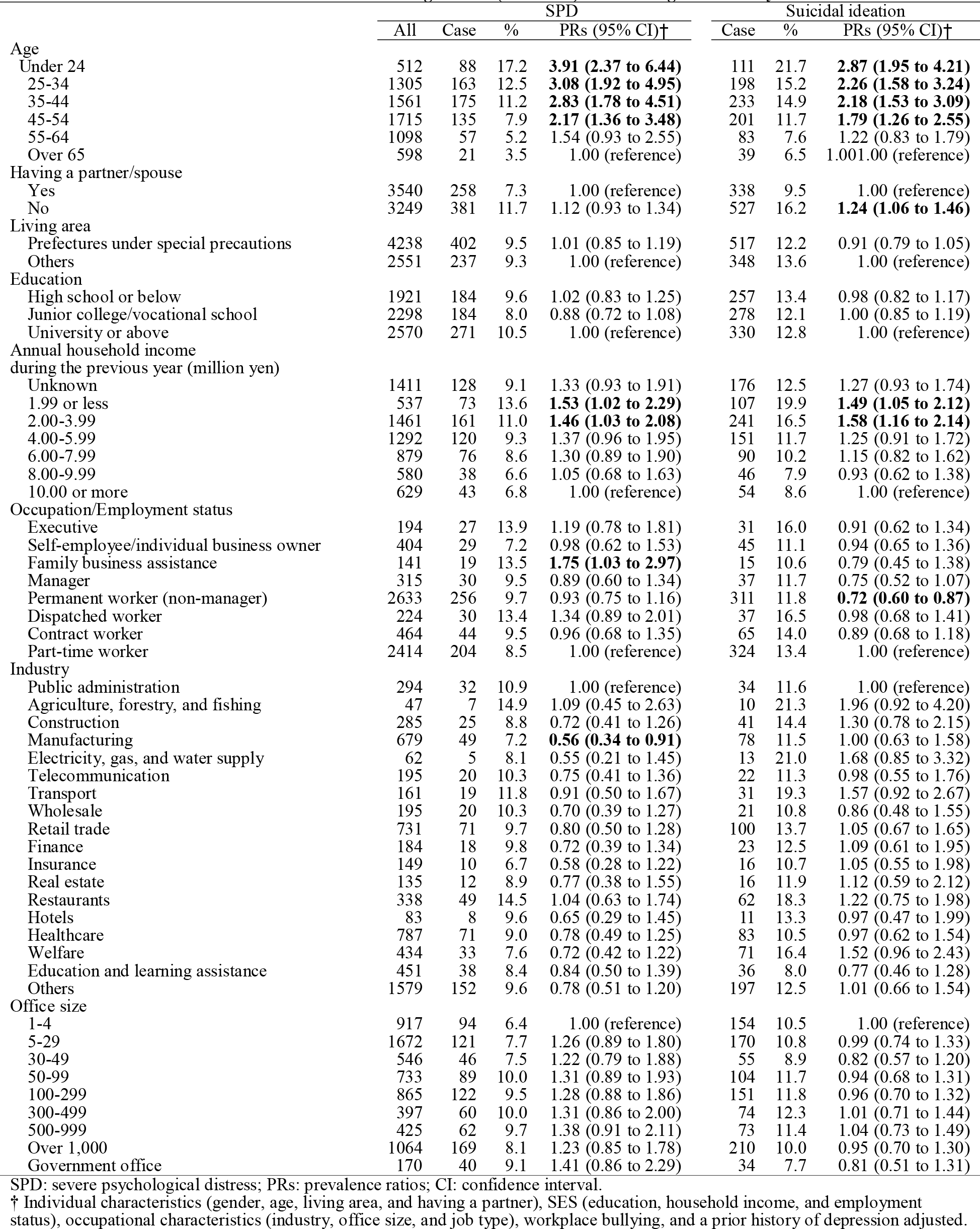
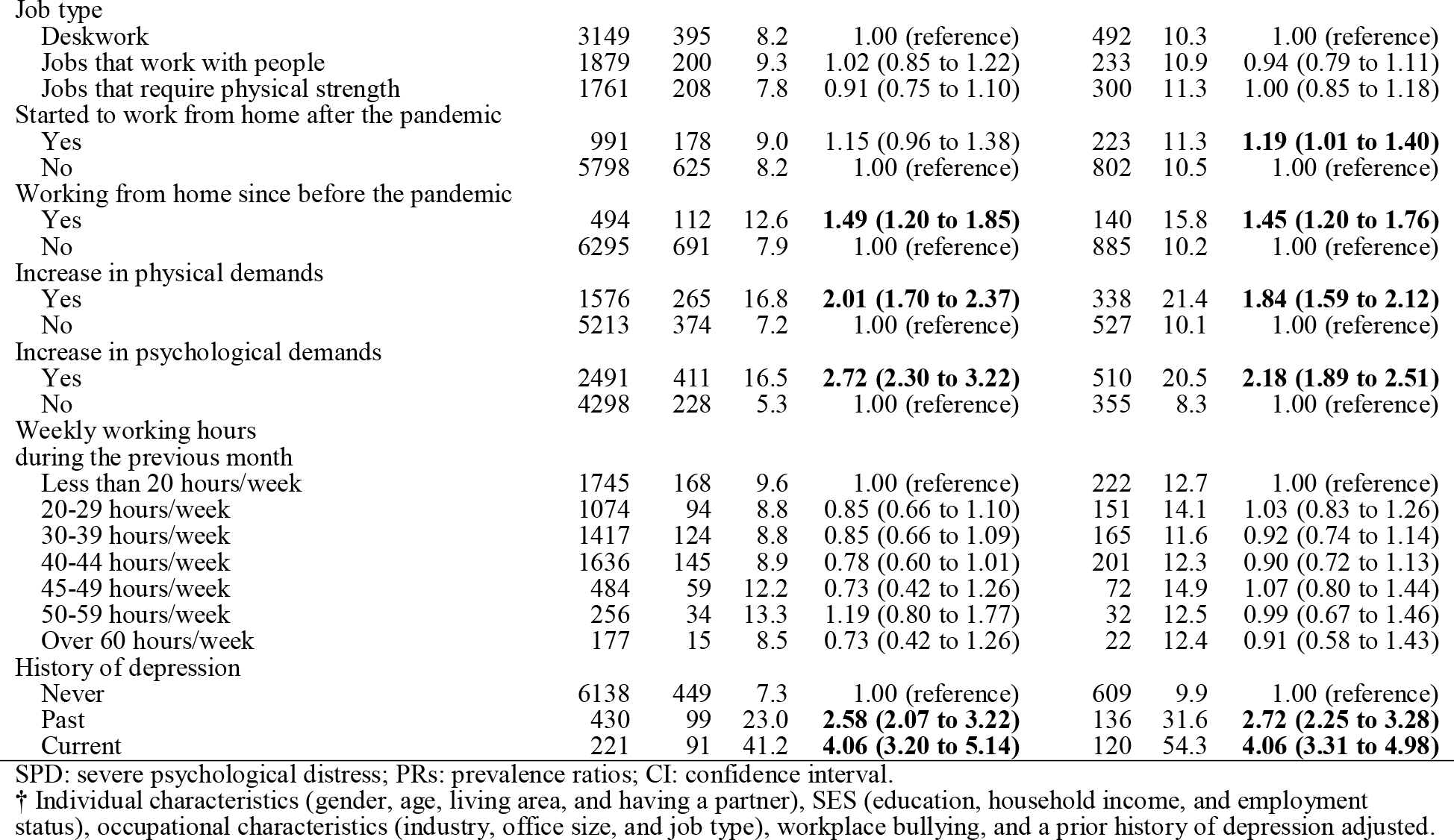
Risk factors for mental health outcomes among women (N = 6.789): Poisson regression analysis

|  | SPD |  |  |  | Suicidal ideation |  |  |
| --- | --- | --- | --- | --- | --- | --- | --- |
|  | All | Case | % | PRs (95% CI)† | Case | % | PRs (95% CI)† |
| Age |  |  |  |  |  |  |  |
| Under 24 | 512 | 88 | 17.2 | **3.91 (2.37 to 6.44)** | 111 | 21.7 | **2.87 (1.95 to 4.21)** |
| 25-34 | 1305 | 163 | 12.5 | **3.08 (1.92 to 4.95)** | 198 | 15.2 | **2.26 (1.58 to 3.24)** |
| 35-44 | 1561 | 175 | 11.2 | **2.83 (1.78 to 4.51)** | 233 | 14.9 | **2.18 (1.53 to 3.09)** |
| 45-54 | 1715 | 135 | 7.9 | **2.17 (1.36 to 3.48)** | 201 | 11.7 | **1.79 (1.26 to 2.55)** |
| 55-64 | 1098 | 57 | 5.2 | 1.54 (0.93 to 2.55) | 83 | 7.6 | 1.22 (0.83 to 1.79) |
| Over 65 | 598 | 21 | 3.5 | 1.00 (reference) | 39 | 6.5 | 1.00 (reference) |
| Having a partner/spouse |  |  |  |  |  |  |  |
| Yes | 3540 | 258 | 7.3 | 1.00 (reference) | 338 | 9.5 | 1.00 (reference) |
| No | 3249 | 381 | 11.7 | 1.12 (0.93 to 1.34) | 527 | 16.2 | **1.24 (1.06 to 1.46)** |
| Living area |  |  |  |  |  |  |  |
| Prefectures under special precautions | 4238 | 402 | 9.5 | 1.01 (0.85 to 1.19) | 517 | 12.2 | 0.91 (0.79 to 1.05) |
| Others | 2551 | 237 | 9.3 | 1.00 (reference) | 348 | 13.6 | 1.00 (reference) |
| Education |  |  |  |  |  |  |  |
| High school or below | 1921 | 184 | 9.6 | 1.02 (0.83 to 1.25) | 257 | 13.4 | 0.98 (0.82 to 1.17) |
| Junior college/vocational school | 2298 | 184 | 8.0 | 0.88 (0.72 to 1.08) | 278 | 12.1 | 1.00 (0.85 to 1.19) |
| University or above | 2570 | 271 | 10.5 | 1.00 (reference) | 330 | 12.8 | 1.00 (reference) |
| Annual household income during the previous year (million yen) |  |  |  |  |  |  |  |
| Unknown | 1411 | 128 | 9.1 | 1.33 (0.93 to 1.91) | 176 | 12.5 | 1.27 (0.93 to 1.74) |
| 1.99 or less | 537 | 73 | 13.6 | **1.53 (1.02 to 2.29)** | 107 | 19.9 | **1.49 (1.05 to 2.12)** |
| 2.00-3.99 | 1461 | 161 | 11.0 | **1.46 (1.03 to 2.08)** | 241 | 16.5 | **1.58 (1.16 to 2.14)** |
| 4.00-5.99 | 1292 | 120 | 9.3 | 1.37 (0.96 to 1.95) | 151 | 11.7 | 1.25 (0.91 to 1.72) |
| 6.00-7.99 | 879 | 76 | 8.6 | 1.30 (0.89 to 1.90) | 90 | 10.2 | 1.15 (0.82 to 1.62) |
| 8.00-9.99 | 580 | 38 | 6.6 | 1.05 (0.68 to 1.63) | 46 | 7.9 | 0.93 (0.62 to 1.38) |
| 10.00 or more | 629 | 43 | 6.8 | 1.00 (reference) | 54 | 8.6 | 1.00 (reference) |
| Occupation/Employment status |  |  |  |  |  |  |  |
| Executive | 194 | 27 | 13.9 | 1.19 (0.78 to 1.81) | 31 | 16.0 | 0.91 (0.62 to 1.34) |
| Self-employee/individual business owner | 404 | 29 | 7.2 | 0.98 (0.62 to 1.53) | 45 | 11.1 | 0.94 (0.65 to 1.36) |
| Family business assistance | 141 | 19 | 13.5 | **1.75 (1.03 to 2.97)** | 15 | 10.6 | 0.79 (0.45 to 1.38) |
| Manager | 315 | 30 | 9.5 | 0.89 (0.60 to 1.34) | 37 | 11.7 | 0.75 (0.52 to 1.07) |
| Permanent worker (non-manager) | 2633 | 256 | 9.7 | 0.93 (0.75 to 1.16) | 311 | 11.8 | **0.72 (0.60 to 0.87)** |
| Dispatched worker | 224 | 30 | 13.4 | 1.34 (0.89 to 2.01) | 37 | 16.5 | 0.98 (0.68 to 1.41) |
| Contract worker | 464 | 44 | 9.5 | 0.96 (0.68 to 1.35) | 65 | 14.0 | 0.89 (0.68 to 1.18) |
| Part-time worker | 2414 | 204 | 8.5 | 1.00 (reference) | 324 | 13.4 | 1.00 (reference) |
| Industry |  |  |  |  |  |  |  |
| Public administration | 294 | 32 | 10.9 | 1.00 (reference) | 34 | 11.6 | 1.00 (reference) |
| Agriculture, forestry, and fishing | 47 | 7 | 14.9 | 1.09 (0.45 to 2.63) | 10 | 21.3 | 1.96 (0.92 to 4.20) |
| Construction | 285 | 25 | 8.8 | 0.72 (0.41 to 1.26) | 41 | 14.4 | 1.30 (0.78 to 2.15) |
| Manufacturing | 679 | 49 | 7.2 | **0.56 (0.34 to 0.91)** | 78 | 11.5 | 1.00 (0.63 to 1.58) |
| Electricity, gas, and water supply | 62 | 5 | 8.1 | 0.55 (0.21 to 1.45) | 13 | 21.0 | 1.68 (0.85 to 3.32) |
| Telecommunication | 195 | 20 | 10.3 | 0.75 (0.41 to 1.36) | 22 | 11.3 | 0.98 (0.55 to 1.76) |
| Transport | 161 | 19 | 11.8 | 0.91 (0.50 to 1.67) | 31 | 19.3 | 1.57 (0.92 to 2.67) |
| Wholesale | 195 | 20 | 10.3 | 0.70 (0.39 to 1.27) | 21 | 10.8 | 0.86 (0.48 to 1.55) |
| Retail trade | 731 | 71 | 9.7 | 0.80 (0.50 to 1.28) | 100 | 13.7 | 1.05 (0.67 to 1.65) |
| Finance | 184 | 18 | 9.8 | 0.72 (0.39 to 1.34) | 23 | 12.5 | 1.09 (0.61 to 1.95) |
| Insurance | 149 | 10 | 6.7 | 0.58 (0.28 to 1.22) | 16 | 10.7 | 1.05 (0.55 to 1.98) |
| Real estate | 135 | 12 | 8.9 | 0.77 (0.38 to 1.55) | 16 | 11.9 | 1.12 (0.59 to 2.12) |
| Restaurants | 338 | 49 | 14.5 | 1.04 (0.63 to 1.74) | 62 | 18.3 | 1.22 (0.75 to 1.98) |
| Hotels | 83 | 8 | 9.6 | 0.65 (0.29 to 1.45) | 11 | 13.3 | 0.97 (0.47 to 1.99) |
| Healthcare | 787 | 71 | 9.0 | 0.78 (0.49 to 1.25) | 83 | 10.5 | 0.97 (0.62 to 1.54) |
| Welfare | 434 | 33 | 7.6 | 0.72 (0.42 to 1.22) | 71 | 16.4 | 1.52 (0.96 to 2.43) |
| Education and learning assistance | 451 | 38 | 8.4 | 0.84 (0.50 to 1.39) | 36 | 8.0 | 0.77 (0.46 to 1.28) |
| Others | 1579 | 152 | 9.6 | 0.78 (0.51 to 1.20) | 197 | 12.5 | 1.01 (0.66 to 1.54) |
| Office size |  |  |  |  |  |  |  |
| 1-4 | 917 | 94 | 6.4 | 1.00 (reference) | 154 | 10.5 | 1.00 (reference) |
| 5-29 | 1672 | 121 | 7.7 | 1.26 (0.89 to 1.80) | 170 | 10.8 | 0.99 (0.74 to 1.33) |
| 30-49 | 546 | 46 | 7.5 | 1.22 (0.79 to 1.88) | 55 | 8.9 | 0.82 (0.57 to 1.20) |
| 50-99 | 733 | 89 | 10.0 | 1.31 (0.89 to 1.93) | 104 | 11.7 | 0.94 (0.68 to 1.31) |
| 100-299 | 865 | 122 | 9.5 | 1.28 (0.88 to 1.86) | 151 | 11.8 | 0.96 (0.70 to 1.32) |
| 300-499 | 397 | 60 | 10.0 | 1.31 (0.86 to 2.00) | 74 | 12.3 | 1.01 (0.71 to 1.44) |
| 500-999 | 425 | 62 | 9.7 | 1.38 (0.91 to 2.11) | 73 | 11.4 | 1.04 (0.73 to 1.49) |
| Over 1,000 | 1064 | 169 | 8.1 | 1.23 (0.85 to 1.78) | 210 | 10.0 | 0.95 (0.70 to 1.30) |
| Government office | 170 | 40 | 9.1 | 1.41 (0.86 to 2.29) | 34 | 7.7 | 0.81 (0.51 to 1.31) |

SPD: severe psychological distress; PRs: prevalence ratios; CI: confidence interval.
† Individual characteristics (gender, age, living area, and having a partner), SES (education, household income, and employment status), occupational characteristics (industry, office size, and job type), workplace bullying, and a prior history of depression adjusted.

**Table 5 (cont.). Risk factors for mental health outcomes among women (N = 6,789): Poisson regression analysis**
|  | SPD |  | Suicidal ideation |
| --- | --- | --- | --- |

|  | All | Case | % | PRs (95% CI)† | Case | % | PRs (95% CI)† |
| --- | --- | --- | --- | --- | --- | --- | --- |
| Job type |  |  |  |  |  |  |  |
| Deskwork | 3149 | 395 | 8.2 | 1.00 (reference) | 492 | 10.3 | 1.00 (reference) |
| Jobs that work with people | 1879 | 200 | 9.3 | 1.02 (0.85 to 1.22) | 233 | 10.9 | 0.94 (0.79 to 1.11) |
| Jobs that require physical strength | 1761 | 208 | 7.8 | 0.91 (0.75 to 1.10) | 300 | 11.3 | 1.00 (0.85 to 1.18) |
| Started to work from home after the pandemic |  |  |  |  |  |  |  |
| Yes | 991 | 178 | 9.0 | 1.15 (0.96 to 1.38) | 223 | 11.3 | **1.19 (1.01 to 1.40)** |
| No | 5798 | 625 | 8.2 | 1.00 (reference) | 802 | 10.5 | 1.00 (reference) |
| Working from home since before the pandemic |  |  |  |  |  |  |  |
| Yes | 494 | 112 | 12.6 | **1.49 (1.20 to 1.85)** | 140 | 15.8 | **1.45 (1.20 to 1.76)** |
| No | 6295 | 691 | 7.9 | 1.00 (reference) | 885 | 10.2 | 1.00 (reference) |
| Increase in physical demands |  |  |  |  |  |  |  |
| Yes | 1576 | 265 | 16.8 | **2.01 (1.70 to 2.37)** | 338 | 21.4 | **1.84 (1.59 to 2.12)** |
| No | 5213 | 374 | 7.2 | 1.00 (reference) | 527 | 10.1 | 1.00 (reference) |
| Increase in psychological demands |  |  |  |  |  |  |  |
| Yes | 2491 | 411 | 16.5 | **2.72 (2.30 to 3.22)** | 510 | 20.5 | **2.18 (1.89 to 2.51)** |
| No | 4298 | 228 | 5.3 | 1.00 (reference) | 355 | 8.3 | 1.00 (reference) |
| Weekly working hours during the previous month |  |  |  |  |  |  |  |
| Less than 20 hours/week | 1745 | 168 | 9.6 | 1.00 (reference) | 222 | 12.7 | 1.00 (reference) |
| 20-29 hours/week | 1074 | 94 | 8.8 | 0.85 (0.66 to 1.10) | 151 | 14.1 | 1.03 (0.83 to 1.26) |
| 30-39 hours/week | 1417 | 124 | 8.8 | 0.85 (0.66 to 1.09) | 165 | 11.6 | 0.92 (0.74 to 1.14) |
| 40-44 hours/week | 1636 | 145 | 8.9 | 0.78 (0.60 to 1.01) | 201 | 12.3 | 0.90 (0.72 to 1.13) |
| 45-49 hours/week | 484 | 59 | 12.2 | 0.73 (0.42 to 1.26) | 72 | 14.9 | 1.07 (0.80 to 1.44) |
| 50-59 hours/week | 256 | 34 | 13.3 | 1.19 (0.80 to 1.77) | 32 | 12.5 | 0.99 (0.67 to 1.46) |
| Over 60 hours/week | 177 | 15 | 8.5 | 0.73 (0.42 to 1.26) | 22 | 12.4 | 0.91 (0.58 to 1.43) |
| History of depression |  |  |  |  |  |  |  |
| Never | 6138 | 449 | 7.3 | 1.00 (reference) | 609 | 9.9 | 1.00 (reference) |
| Past | 430 | 99 | 23.0 | **2.58 (2.07 to 3.22)** | 136 | 31.6 | **2.72 (2.25 to 3.28)** |
| Current | 221 | 91 | 41.2 | **4.06 (3.20 to 5.14)** | 120 | 54.3 | **4.06 (3.31 to 4.98)** |

SPD: severe psychological distress; PRs: prevalence ratios; CI: confidence interval.
† Individual characteristics (gender, age, living area, and having a partner), SES (education, household income, and employment status), occupational characteristics (industry, office size, and job type), workplace bullying, and a prior history of depression adjusted.

## DISCUSSION

In this nationwide internet survey for the general working population, 15% of workers experienced workplace bullying, 9% had SPD, and 12% had suicidal ideation during the second and third wave of the COVID-19 pandemic in Japan (April-September 2020). The results of this study showed younger age, low household income, increase in physical demands, increase in psychological demands, and a prior history of depression were common significant risk factors for workplace bullying, SPD, and suicidal ideation. Although this pattern is similar to the trend before the pandemic,^6 8^ a different pattern was also observed in this study: men and workers with higher occupational positions such as executives, managers, or permanent workers had a higher risk of bullying compared to women or part-time workers. As job workload was reported as antecedents of bullying,^6 13–15^ the COVID-19-related working environment changes, such as an increase in physical or psychological demands, may affect the findings. A new working style—working from home—was also associated with adverse mental health, however, starting working from home was found to be a preventive factor against workplace bullying. This indicates that working from home has both advantages and drawbacks; although starting working from home contributes to decreasing aggressive and negative acts from supervisors or co-workers, it makes workers being isolated due to lack of mutual communication between work members.^24^ This may contribute to having psychological distress because the amount of worksite social support decreased at the same time.^23^ Overall, the results of this study implicate that when intervening to prevent workplace bullying or mental health problems among workers, we should focus on not only previously reported vulnerable workers but also workers who experienced a change of their working styles or their job demands.

The prevalence of workplace bullying in this study was similar to the global prevalence before the pandemic.^1^ Although it was higher than previously reported in the representative working sample in Japan (6.1%),^8^ this does not immediately mean that more workers experience bullying at work during the pandemic because the measurement durations are different; the previous study measured experiencing workplace bullying during the month but this study measured during the previous six months. As previously reported, measurement methods greatly contributed to the prevalence rates of workplace bullying.^1^ As a recent national survey of workplace bullying and harassment in Japan showed a non-different prevalence of workplace bullying in 2020 (31.4%) and in 2016 (32.5%), the prevalence itself may not be changed before and during the pandemic in Japan.

In this study, men, younger workers, workers with lower household income, executives, managers, permanent workers, contract workers, workers working in larger office size, workers experienced an increase in physical or psychological demands, and workers with current or a prior history of depression or other mental illnesses were more likely to expose to workplace bullying. Although most of the results are consistent with previous studies,^2 5 6 8 13 14^ inconsistent results were observed in terms of gender and occupational positions in this study;^9^ men, executives, and managers had a higher risk of experiencing workplace bullying compared to women and part-time workers. This trend may be caused by the pandemic since an increase in physical or psychological demands was also associated with exposure to bullying in this study. During the pandemic, managers had to determine what countermeasures should be implemented to protect employees against COVID-19. At the same time, managers had to follow the government statements or guidelines against COVID-19, which may decrease their job autonomy or control.^14^ Moreover, during the pandemic, executives and managers had to adapt new technologies such as online meetings or new working styles including working from home.^18^ Taking into account that most of the executives and managers probably did not have expertise in infection control or new technologies, this may boost subordinates’ frustration, which may cause aggressive behaviors toward managers.^32^ A study for managers has reported the risk for a manager to being bullied was higher in those who suffer from work stress, those less satisfied with their payment, and those who do not see opportunities for promoting within their organizations.^10^ Managers in Japan have been reported as highly stressful workers since most of them are middle managers with heavy workloads and limited autonomy, who are often described as ”player managers” (managers who have double burdens: players and managers).^33^ This also may contribute to a high prevalence of workplace bullying among managers in this study.

Exposure to workplace bullying was significantly associated with SPD and suicidal ideation both in men and women, even after adjusting for individual characteristics, SES, occupational characteristics, and a prior history of depression. This indicates a strong relationship between bullying and mental health problems, as previously shown.^2–4^ ^34^ Moreover, not exposed but witnessed bullying was also associated with both SPD and suicidal ideation in this study. This is in line with the longitudinal study which showed a spillover effect of workplace bullying on psychological distress.^34^ Our findings indicate that this effect could be applied to workers’ suicidal ideation.

Men were more likely to have SPD or suicidal ideation than women when being bullied. Although gender differences have not been investigated in the meta-analyses on the association between bullying and mental health,^2–4^ the results of this study are consistent with the study of the association between work-related physical violence and depression in Japan.^35^ Two possible explanations are considered: first, men were more likely to be in managerial positions than women. Since a high prevalence of workplace bullying was observed in executives and managers in this study, this may affect the high prevalence of SPD or suicidal ideation in men. The second possible explanation is low psychological preparedness, which refers to a sense of control over the trauma.^36^ Since men had a lower risk of workplace bullying before the pandemic, men who experienced bullying may tend to feel shocked and have more severe mental health problems than women who have a higher risk for bullying in general.^6 7^

Newly starting working from home was a preventive factor against workplace bullying, while it was a predictor for adverse mental health outcomes in this study. This indicates that working from home has both advantages and drawbacks; although newly introduced working from home decreases communication among workers including aggressive and negative acts from supervisors or co-workers, it makes workers being isolated from co-workers. Thus, this may contribute to workers’ deterioration of mental health because the amount of worksite social support was decreased at the same time.^23^ In this study, although working from home since before the pandemic was associated with both SPD and suicidal ideation, starting to work from home during the pandemic was associated with only suicidal ideation. This is consistent with the study reported long-term working from home hindered workers’ mutual communication or supports from co-workers.^23^ ^24^ Thus, the results of this study implicate that working from home contributes to protecting workers from negative acts, but the amount of worksite social support should not be decreased and workers’ mental health status should be monitored regularly for remote workers.

We found younger age, lowest household income, increase in physical demands, increase in psychological demands, and a history of depression were risk factors of SPD and suicidal ideation both in men and women, in addition to workplace bullying and working from home. Although existing literature already showed the significant association between job demands and mental health,^37^ this study added to it that changes in job demands may also affect SPD or suicidal ideation during the pandemic. In contrast, the absence of a partner/spouse was associated with SPD and suicidal ideation in men but only associated with suicidal ideation in women. This is probably because the benefit of having a partner/husband was greater in men than women on mental health and having a partner/husband does not mean she can gain enough amount of support from him.^38^

Several limitations should be noted. First, this study was cross-sectional so that causality cannot be determined. Although we adjusted for a prior history of depression to omit reverse causality, a longitudinal study is needed to clarify the association between risk factors and workplace bullying, SPD, and suicidal ideation in the COVID-19 pandemic. Second, workplace bullying was measured by a self-labeling method, which may cause underestimation compared to the behavioral experience method that asks respondents how often they experienced the various negative acts without using the term “harassment” or “bullying.”^1^ Third, there might be a sampling bias due to the nature of an online survey. This may limit generalizability of our study results.

Despite these limitations, this study was the first to identify important risk factors of workplace bullying, SPD, and suicidal ideation in the nationwide large-scale study for the general working population in Japan. The strengths of this study were investigating various risk factors including working styles and a change in job demands and revealed new risk factors: working from home for SPD or suicidal ideation and managers for workplace bullying. Further research is needed to examine other possible risk factors of workplace bullying, SPD, and suicidal ideation.

## CONCLUSIONS

Overall, 15% of workers experienced workplace bullying, 9% had SPD, and 12% had suicidal ideation during the second and third wave of the COVID-19 pandemic in Japan. The results of this study showed men, executives, managers, and permanent workers had a higher risk of bullying compared to women or part-time workers. Workers who experienced increase in physical or psychological demands were at risk for bullying, SPD, and suicidal ideation. Newly starting working from home was a significant predictor for adverse mental health outcomes, however, it was found to be a preventive factor against workplace bullying. The results of this study show a different pattern of high-risk groups during the pandemic. When intervening to decrease workplace bullying or mental health problems, we should focus on not only previously reported vulnerable workers but also new high-risk groups or workers who experienced a change of their working styles or job demands.

## Data Availability

The data used in this study are not available in a public repository because they contain personally identifiable or potentially sensitive patient information. Based on the regulations for ethical guidelines in Japan, the Research Ethics Committee of the Osaka International Cancer Institute has imposed restrictions on the dissemination of the data collected in this study. All data inquiries should be addressed to the person responsible for data management, Dr. Takahiro Tabuchi at the following e-mail address.

## Acknowledgments

We are grateful to other research members of JACSIS for their tremendous efforts to conduct this survey.

## Contributors

KT conceived the research idea. KT and TT conducted a survey and gathered the data. KT carried out the statistical work and wrote the first draft. KT and TT revised it and wrote the final version of the paper.

## Patient consent for publication

Not required.

## Ethical approval

The study protocol was reviewed and approved by the Research Ethics Committee of the Osaka International Cancer Institute (approved on June 19, 2020; approval number 20084). The internet survey agency respected the Act on the Protection of Personal Information in Japan. All participants provided web-based informed consent before responding to the online questionnaire. A credit point known as “Epoints”, which could be used for internet shopping and cash conversion, was provided to the participants as an incentive.

## Provenance and peer review

Not commissioned; externally peer reviewed.

